# Maintaining EHR-based Predictive Clinical Decision Support in Mental Health against Temporal Shifts

**DOI:** 10.64898/2026.10.28.26366683

**Authors:** Xin Xiong, Elliott Shi, Zhouhan Qian, Haobo Zhu, Chuan Hong, Zijian Guo, Jordan W Smoller, Molei Liu, Tianxi Cai

**Author notes:** these authors jointly supervised the project (Jordan W Smoller, Molei Liu, Tianxi Cai).

## Abstract

Electronic health record (EHR)-based risk prediction offers opportunities for early intervention in mental health, but models may remain in use as patient populations, care practices, and documentation change. Such temporal shifts can reduce predictive performance, while updating models from recent data alone may be unstable when adverse outcomes are uncommon. We developed Adversarial Drift-Aware Predictive Transfer (ADAPT), a distributionally robust approach to improve the temporal durability of mental health risk prediction. ADAPT uses variation in models estimated from previous calendar periods to characterize plausible changes in predictor–outcome relationships. Data from the current period constrain which historical patterns remain relevant, allowing the method to seek robustness against subsequent shifts while preserving contemporary discrimination. We retrospectively evaluated ADAPT using EHRs from 889,445 patients with mental health diagnoses at Mass General Brigham and Duke University Health System, covering one-year risk prediction for 28 outcomes. In the worst 20% of year-pair comparisons, ADAPT improved AUC over all comparators by 0.016–0.036 at Mass General Brigham, 0.011–0.018 at Duke, and 0.008–0.037 after transport. For suicide-attempt prediction, contemporary AUC at Mass General Brigham was 0.82 compared with 0.80 for the best comparator. Eight-year relative AUC degradation was 7.7%, compared with 12.1% for pooled-data training. These results suggest that information from prior temporal shifts can improve robustness to future changes without sacrificing predictive performance for current patients.

## 1 Introduction

Clinical AI is increasingly moving from research towards clinical implementation. This transition is reflected in the increasing number of machine-learning (ML) and AI-enabled clinical technologies authorized by the US Food and Drug Administration and in the adoption of predictive AI within health systems (Shick et al., 2024). In a 2024 national survey of US hospitals, more than half reported using predictive AI integrated with their electronic health record (EHR) systems (Everson et al., 2025). As AI models become embedded within EHR systems and integrated into clinical workflows to support clinical decision making, attention must extend beyond their performance at initial development and validation to their reliability after deployment. Models may remain in use for years while the patients, care practices, technologies, and health-care environments around them continue to change. Maintaining predictive performance over time is therefore becoming an increasingly important component of safe and effective clinical AI.

Mental health provides a particularly important clinical context to study this problem. Mental disorders impose a substantial and growing burden on patients, families, health systems, and society and contribute extensively to global disability (GBD 2023 Mental Disorder Collaborators, 2026). Because treatment alone has not adequately reduced this burden, prevention and early intervention have become central priorities, including strategies that identify individuals at elevated risk before serious adverse outcomes occur (Arango et al., 2018). Risk prediction models using longitudinal EHR data offer an opportunity to support these goals by identifying patients who may benefit from earlier intervention. EHR-based prediction models have been developed for antidepressant treatment response, psychiatric readmission, mental health crises, self-harm, and suicide attempts (Sheu et al., 2023a; Ren et al., 2025; Garriga et al., 2022; Barak-Corren et al., 2017; Simon et al., 2018; Sheu et al., 2023b). Importantly, some of these models have progressed beyond retrospective development to prospective or trial-based evaluation within clinical workflows, highlighting their potential use as clinical decision-support tools (Garriga et al., 2022; Walsh et al., 2025). As such models move closer to routine clinical use, ensuring that their performance remains reliable after deployment becomes increasingly important.

Mental health may also be particularly susceptible to temporal shift because both the underlying risk and its representation in EHR data can be influenced by social, environmental, and health-system conditions (Choudhary et al., 2025). Changes in insurance coverage, access to care, treatment availability, screening practice, documentation, substance use, and patterns of healthcare utilization can alter both who is observed within a health system, how risk factors and outcomes are captured, and the distribution of the predictors themselves. Major external disruptions such as COVID-19 can affect several of these mechanisms simultaneously. During the COVID-19 pandemic, for example, primary-care-recorded self-harm changed abruptly and differently across patient subgroups, with observed reductions potentially reflecting changes in help-seeking as well as changes in underlying incidence (Steeg et al., 2021; Reger et al., 2020). Changes in policy and EHR systems can have similar disruptions. Implementation of universal suicide screening in emergency departments nearly doubled documented detection of suicide risk (Boudreaux et al., 2016), while transitions between coding systems, including the change from ICD-9 to ICD-10, can alter the granularity and comparability of recorded diagnoses, injuries, and mental-health-related outcomes (Mainor et al., 2019; Cartwright, 2013). Mental health therefore provides a clinically consequential setting in which changes in the external environment, clinical practice, and EHR data generation can occur together.

For a deployed AI model, these changes can produce data shift, a mismatch between the data on which a model was developed and the post-deployment data, which is an important cause of the failure to perform reliably (Finlayson et al., 2021). As patient populations, treatments, screening practices, coding, documentation, access to care, and clinical workflows evolve, models trained using older data may become progressively less reliable. Such degradation can directly undermine clinical decision support: false-negative predictions may create missed opportunities for intervention, whereas false-positive alerts can increase screening and follow-up burden and divert limited clinical resources. Temporal degradation has been observed across clinical AI applications and can emerge within only a few years of model development (Vela et al., 2022; Fernández-Narro et al., 2025).

As such, models intended for longitudinal clinical use must not only demonstrate strong performance at initial development and validation, but also maintain their accuracy across changing clinical environments and over time. This creates an additional challenge beyond initial model development and validation: how to maintain predictive performance once a model is deployed (Feng et al., 2022). In practice, clinical prediction models have often been treated as static tools, with updating performed periodically, ad hoc, or after evidence of performance deterioration has accumulated (Jenkins et al., 2021). Maintaining performance through repeated updating can itself be difficult: reliable outcome labels may not become available promptly, and model changes require continued performance assessment, validation, and governance (Davis et al., 2024; Subasri et al., 2025).

Common model-updating strategies also involve important trade-offs. Pooling all historical observations can improve precision but may dilute recent changes, whereas fitting a model using only the most recent period can be unstable especially when the contemporary sample contains few adverse outcomes. Transfer-learning and domain-adaptation methods can borrow information from historical data, but they generally focus on performance in a known current target population rather than durability under subsequent shifts not yet observed (Gretton et al., 2009; Reddi et al., 2015; Liu et al., 2023; Li et al., 2022; Tian and Feng, 2023). Therefore, a complementary need is methods that can use variations across prior data sources to improve robustness to potential future shifts while preserving strong performance in the observed training target data.

To address this need, we developed Adversarial Drift-Aware Predictive Transfer (ADAPT), an approach designed to improve the temporal durability of EHR-based prediction models while preserving performance for contemporary patients. ADAPT uses variations across model estimates from prior calendar periods, together with limited data from the most recent patient population, within a distributionally robust optimization (DRO) framework to guide model development toward robustness against potential future shifts. ADAPT leverages how the predictor-outcome relationships have varied across earlier temporal periods while using contemporary data to constrain the influence of historical patterns that may no longer be relevant. As such, ADAPT balances strong performance for patients seen today with reduced vulnerability to changes that may occur after deployment.

ADAPT was also designed to facilitate implementation across health systems. Historical calendar periods contribute through summary-level fitted-model estimates without requiring historical patient-level data to be repeatedly pooled or transferred, and only limited contemporary data are needed to anchor the model to the current population. The resulting model retains the same predictor structure as earlier models and can thus be applied to new patients using the same architecture. This reduces the data-sharing and computational requirements associated with model maintenance.

We examined whether ADAPT could support more durable mental health risk assessment. Using longitudinal EHR data from Mass General Brigham and Duke University Health System, we retrospectively evaluated one-year risk across 28 mental health outcomes, both within and across health systems. Our evaluation spanned both the transition from ICD-9 to ICD-10 coding and the COVID-19 pandemic, testing whether models developed in an earlier year remained reliable for future patients through major changes in clinical documentation and care delivery. ADAPT improved discrimination across a range of conditions, including psychotic disorders, bipolar disorder, and suicidal ideation, with gains extending to the periods in which predictive performance was poorest. In a focused suicide-attempt case study, ADAPT combined strong current-year discrimination with reduced deterioration when the same fitted model was applied to patients seen years later, and this advantage was retained after transfer to another health system. Together, these findings support the potential for greater model longevity, i.e., preserving predictive performance beyond the year of development while maintaining accuracy for contemporary patients, and for mental health decision-support tools that remain informative as patient populations and care practices evolve.

## 2 Methods

### 2.1 Study design and data sources

We conducted a retrospective, multi-health-system study to evaluate the temporal durability and cross-institutional transportability of EHR-based risk prediction models for a range of mental health outcomes. We evaluated ADAPT independently at both Mass General Brigham (MGB) and Duke University Health System (Duke), developing models sequentially across calendar years and assessing contemporaneous performance and temporal degradation within each health system. We additionally applied models developed at MGB directly to Duke data across calendar years to assess cross-institution transportability. We conducted two complementary analyses across both institutions: 1) a multi-outcome analysis evaluated ADAPT across 28 mental-health outcomes; and 2) a suicide-attempt case study focused on incident suicide attempts. Unless otherwise specified, both analyses used the same predictor construction, model-development, and validation procedures.

The MGB EHR cohort was obtained from its Research Patient Data Registry, a centralised EHR repository containing records for more than 7 million patients across multiple affiliated hospitals, including Massachusetts General Hospital and Brigham and Women’s Hospital (Sheu et al., 2023a). The MGB cohort included records from Jan 1, 2005, to Dec 31, 2022. The Duke database contains longitudinal EHR records for more than 6 million patients with records spanning from Jan 1, 2014 to Dec 31, 2022. The same general cohort-construction, outcome-definition, feature-generation, and model-evaluation procedures were applied at both institutions.

### 2.2 Study population, outcome, and predictors

#### Multi-outcome analysis

Patients were eligible if they had at least one recorded diagnosis of mental health-related condition, defined using PheCodes 295–318, and at least six months of follow-up. PheCodes group related ICD codes into categories to represent clinically meaningful broader clinical phenotypes such as PheCode 296.22 for major depressive disorder and 297.1 for suicide ideation (Denny et al., 2013). The landmark visit was defined as the recorded visit closest to one year after the first mental health diagnosis, and patients with less than one month of follow-up after the landmark visit were excluded. We evaluated ADAPT across 28 mental-health outcomes, selected as the subset of PheCodes 295–318 with at least 15 cases within one year of the landmark visit at both MGB and Duke. Each outcome was treated as a separate prediction task, with the objective of predicting its occurrence within one year after the landmark visit. Outcome definitions were harmonised across MGB and Duke using corresponding PheCodes. For each outcome, we focused on patients *without* a prior diagnosis of that outcome. Predictors were measured during the three years preceding the landmark visit.

#### Suicide-attempt case study

The suicide-attempt case study used a separate landmark definition. Following Sheu et al. (2023b), the landmark date was randomly selected from the patient’s recorded visits between the first mental health diagnosis and the 75th percentile of the visit dates after that diagnosis. We defined an incident suicide attempt as an event occurring within one year after the landmark visit and identified attempts using the ICD-9-CM and ICD-10-CM code lists from two previous EHR-based suicide-prediction studies (Barak-Corren et al., 2020, 2017). Predictors were measured from one year before the landmark visit.

#### Predictors

For each outcome, candidate predictors were selected from the codified feature list provided by the Online Narrative and Codified feature Extraction (ONCE) pipeline, a knowledge-graph-based approach that represents EHR features and quantifies their relative importance for each target PheCode (Xiong et al., 2023). Raw EHR codes were mapped to standard ontologies and included diagnoses represented by PheCodes, medications by RxNorm, procedures by Clinical Classifications Software categories, and laboratory tests represented by LOINC (Huang et al., 2025). Each codified feature was represented as its total number of occurrences during the three-year period preceding the landmark visit, transformed as log(1+*x*). The same candidate-feature construction approach was used in both multi-outcome and suicide attempt analyses, with only the feature-counting window differing. Age at the landmark visit, gender, and race were also included as covariates. We applied the same feature generation strategy across prediction tasks to facilitate comparisons across settings.

### 2.3 ADAPT framework

ADAPT was designed to maintain accurate mental-health risk prediction for patients in the current period while reducing deterioration when the same model is applied to subsequent years. For a development year *t*, all earlier *L* calendar periods were treated as historical sources and year *t* as the current target population, indexed by *Q*. For each mental health outcome, a period-specific model took the logistic form

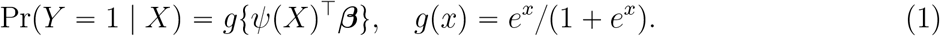

where *Y* indicates the outcome within one year, *β* is the period-specific coefficient vector, and *ψ*(*X*) is a general feature map that includes an intercept and can incorporate different representations of the observed data. These can include original clinical features, latent representation learned from pre-trained models such as transformers or nonlinear features derived from pre-trained machine learning prediction models.

ADAPT proceeds in three steps. In Step 1, the historical-period data was fitted using the model defined above with weighted penalized logistic regression, generating coefficient estimates 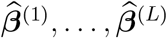. The current-period data are split into two equally sized subsets, 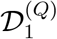 and 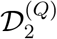. The first subset was used to estimate the current-period coefficient 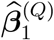 , while the second was held out for constructing the set of potential future models in Step 2. Inverse probability of censoring weighting (Uno et al., 2007) was used in model fitting and evaluation to account for incomplete one-year follow-up.

In Step 2, ADAPT used the historical and current-period models to define a range of potential future predictor–outcome relationships. The construction was guided by two principles. First, variation observed across previous calendar periods provided information about how the predictor–outcome relationship might change in the future, which we represented through mixtures of the historical and current-period models. Second, because the current population was the most relevant reference for near term prediction, we required these mixtures to remain sufficiently compatible with the held-out current-period data. Specifically, collecting the candidate directions as 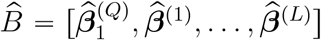 represented all mixtures of the observed predictor–outcome relationships, where Δ_*L*+1_ denotes the set of non-negative weights summing to one. ADAPT then used the held-out current-period data to remove mixtures that were insufficiently compatible with contemporary patients. We therefore defined the constrained set of models representing *emulated future distributions* as

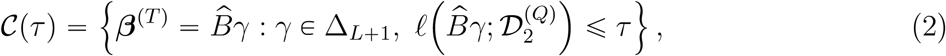

where *l*(·; *D*) is the weighted negative logistic log-likelihood on data *D*. The loss constraint retained mixtures that remained sufficiently compatible with the held-out current-period data.

The tuning parameter *τ* controlled how tightly the candidate set was anchored to the current population: smaller values produced a tighter set, while larger values allowed greater influence from historical variation. The choice of *τ* is described in Supplementary Methods, Section A.2.

In Step 3, ADAPT used distributionally robust optimization to select a model that performed well even under the least favorable relationship within this constrained set:

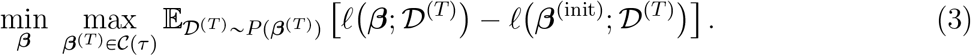

Here *P* (***β***^(*T*)^) denotes a hypothetical future population that retains the predictor distribution of the current period but follows the outcome relationship indexed by ***β***^(*T*)^, and *D*^(*T*)^ denotes data drawn from that population. The reference model ***β***^(init)^ is a user-specified initial guess of the future model. Throughout this paper, we used the sample-size-weighted average of the period-specific estimates 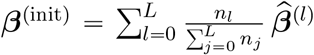. The inner maximization identified the least favorable relationship within *C*(*τ*), and the outer minimization selected the coefficients that made this worst-case excess loss as small as possible. Thus, ADAPT used historical variation to protect against potential future shifts while the current-period constraint limited protection against outdated patterns. By restricting the candidate set using current-period data, ADAPT focused robustness on historical variation that remained relevant to the contemporary population. Our framework differs from transfer learning, which primarily uses historical information to improve estimation in the current observed target population. It also differs from conventional maximin learning, which protects against all mixtures of the observed populations and tends to be overly conservative due to the inclusion of outdated patterns.

Solving the min–max problem in (3) directly is computationally demanding. We thus used a second-order approximation around ***β***^(init)^, reducing the problem to the constrained quadratic optimization

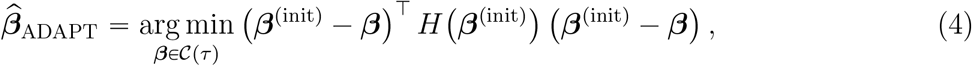

where *H*(***β***^(init)^) is the Hessian of the weighted negative log-likelihood evaluated at ***β***^(init)^. Full derivations and computational details are provided in Supplementary Methods, Section A.2. A practical advantage of ADAPT is that historical periods contribute through summary-level fitted-model estimates rather than pooled historical patient-level records. This reduces the need to repeatedly centralize or transfer historical data and facilitates implementation across institutions when model estimates, but not patient-level data, can be shared.

### 2.4 Validation Studies

#### 2.4.1 Benchmark Methods

We compared ADAPT with four benchmark methods representing common strategies for multi-source model development. The current-period-only model used only the most recent data and was fitted as an *ℓ*_1_-regularized logistic regression only to data from year *t*. The cumulative pooled-data model combined all patient-level data available up to and including year *t* and fitted the same *ℓ*_1_-regularized logistic regression to the pooled data. TransGLM borrowed information from historical periods while adapting the model to the current target population by shrinking the target estimate toward similar source models (Tian and Feng, 2023). As a robustness benchmark, the conventional maximin estimator considered all possible mixtures of the observed source and current target populations and optimized against the worst-performing mixture, without restricting the mixture to remain close to the current target population (Meinshausen and Bühlmann, 2015). Because the original maximin formulation was developed for squared-error regression, it was adapted to logistic regression using a second-order local approximation to the negative log-likelihood around the current target; the resulting quadratic objective used a simplex constraint and a zero-vector initial model. Each comparator was applied separately to each mental-health outcome and within each health-system analysis.

##### Sensitivity analysis with non-linear feature representations

As a sensitivity analysis, we assessed whether incorporating non-linear feature representations changed the relative performance of ADAPT and the benchmark methods. We trained source-specific XGBoost models and appended their prediction scores to the original EHR features. For each calendar year, data were split into a held-out evaluation set (one half), an XGBoost training set (one quarter), and a regression set (one quarter) used to fit all regression-based models. Each year-specific XGBoost model was then applied to the regression and evaluation sets, other source years, and the full target-year data to generate prediction scores. Thus, the augmented features used to fit and evaluate the regression-based models were not derived from in-sample data, thereby avoiding overfitting. We compared ADAPT and the benchmark methods using both the original and augmented feature sets. Details are provided in Supplementary Methods, Section A.3.

##### Temporal and cross-health-system validation

The target year *t* was separately set as 2010, … , 2022 in MGB and *t* ∈ {2015, … , 2022} in Duke, with all years before *t* used as historical sources. Three sets of validation analyses were conducted. First, contemporaneous internal validation was performed separately at MGB and Duke, with models trained using part of the observed target data and validated on the remainder target data within the same calendar year. Second, temporal validation was performed separately at both institutions by applying models developed for year *t* to data from subsequent calendar years within the same health system. Third, to assess cross-institution transportability, models developed at MGB for year *t* were applied directly to Duke data from the calendar year closest to *t* and all the subsequent years. No data from calendar years after the model-development year were used to fit the historical or current-period models. Each target-year dataset was randomly split 10 times, and performance was averaged across repetitions.

##### Synthetic data validation

We additionally conducted a synthetic-data validation to evaluate ADAPT under controlled temporal-shifts. We generated 15 sequential periods in which predictor-outcome relationships evolved gradually over time, with an additional abrupt perturbation introduced at period 8. We varied the amount of current-period data relative to historical data and the magnitude of the abrupt shift, allowing us to assess ADAPT under both limited contemporary data and different degrees of temporal change. Full data-generating details are provided in Supplementary Methods, Section A.4.

### 2.5 Evaluation Metrics

#### Performance metrics for mental-health outcomes

For each mental-health outcome, prediction method, target year *t*, validation year *v* ≥ *t*, we evaluated area under the receiver operating characteristic curve (AUC), sensitivity, and lift. Lift was defined as precision divided by outcome prevalence, representing enrichment of positive cases relative to random selection. For sensitivity and lift, a method- and target-year-specific threshold was selected using the held-out target-year validation data to achieve 0.95 specificity, or the closest attainable value. The threshold was held fixed when the model developed for year *t* was evaluated in later calendar years *v* > *t*. For cross-health-system validation, the threshold selected at MGB was transported with the fitted model and applied unchanged at Duke. For all temporal and cross-health-system validation, model coefficients were likewise held fixed and applied without refitting to each validation cohort.

#### Worst-fraction performance across training–validation year pairs

To summarize robustness across training–validation year pairs, we calculated lower-tail Conditional Value at Risk (CVaR) (Rockafellar and Uryasev, 2000) separately for each mental-health outcome, method, evaluation metric, and validation setting:

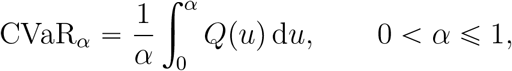

where *Q*(*u*) denotes the empirical performance quantile across eligible training–validation year pairs. CVaR_*α*_ therefore represents the mean performance among the worst-performing fraction *α* of pairs. Smaller values of *α* place greater emphasis on the most difficult temporal settings. As *α* approaches zero, CVaR_*α*_ approaches the worst-pair performance, while CVaR_1_ equals the mean performance across all pairs. The higher CVaR on the lower-tail indicates greater robustness in settings where performance is most vulnerable to degradation. We evaluated CVaR over a prespecified grid of *α* values. For each *α*, CVaR_*α*_ was averaged between PheCodes and ADAPT was compared to each benchmark using differences between methods in mean CVaR_*α*_. Corresponding *p*-values were calculated based on 500 bootstrap replicates. All CVaR analyses were performed separately for the within-MGB, within-Duke, and MGB-to-Duke validation settings.

#### Model aging effect

For the suicide-attempt case study, we further quantified model aging effect by comparing each method’s aged model with its “newborn” model. For a validation year *t*, the newborn model was developed in year *t*, whereas the aged model was developed Δ years earlier; both were evaluated on the same held-out validation data from year *t*. Relative temporal degradation was defined as

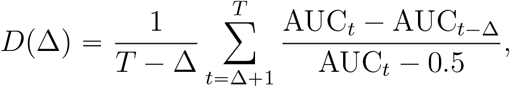

where *t* indexes the *T* study years, and AUC_*t*_ and AUC_*t*−Δ_ denote the AUCs of the newborn and aged models, respectively, evaluated in the same year *t*. This measure expressed the loss in discrimination from the aged model relative to the discrimination of the newborn model above chance, with smaller values indicating less temporal degradation. We calculated *D*(Δ) across increasing training–validation year gaps Δ to compare the rate of temporal degradation between ADAPT and the benchmark methods. For external validation, both models were developed at MGB and evaluated on Duke data from the validation year *t*.

## 3 Results

### 3.1 Study population and temporal patterns

The study included 889,445 patients with mental health diagnoses: 412,943 from MGB and 476,502 from Duke (Table 1). Females comprised 64.5% of the MGB cohort and 56.6% of the Duke cohort. The cohorts differed in racial composition: 6.0% of MGB patients and 27.3% of Duke patients were Black, whereas 79.6% and 64.0%, respectively, were White. Patients aged 26–55 years represented the largest age group at both sites. The full distribution of landmark years is provided in Supplementary Table S1.

**Table 1:** Selected characteristics of the MGB and Duke cohorts. Values are *n* (%).

| | Duke ($N = 476,502$) | MGB ($N = 412,943$) | Overall ($N = 889,445$) |
| --- | --- | --- | --- |
| Female | 269477 (56.6%) | 266318 (64.5%) | 535795 (60.2%) |
| Male | 207020 (43.4%) | 146625 (35.5%) | 353645 (39.8%) |
| Unknown gender | 5 (0.0%) | 0 (0%) | 5 (0.0%) |
| Asian | 10990 (2.3%) | 11008 (2.7%) | 21998 (2.5%) |
| Black | 129915 (27.3%) | 24682 (6.0%) | 154597 (17.4%) |
| White | 305195 (64.0%) | 328755 (79.6%) | 633950 (71.3%) |
| Other race | 17318 (3.6%) | 47367 (11.5%) | 64685 (7.3%) |
| Unknown race | 13084 (2.7%) | 1131 (0.3%) | 14215 (1.6%) |
| Age 0–18 years | 73749 (15.5%) | 27872 (6.7%) | 101621 (11.4%) |
| Age 19–25 years | 34920 (7.3%) | 35919 (8.7%) | 70839 (8.0%) |
| Age 26–55 years | 187999 (39.5%) | 193528 (46.9%) | 381527 (42.9%) |
| Age > 55 years | 179834 (37.7%) | 155624 (37.7%) | 335458 (37.7%) |

### 3.2 Performance across mental-health outcomes

Across the 28 mental-health outcomes, ADAPT generally maintained stronger performance in the more challenging temporal settings. When performance was summarized over the worst-performing half of training–validation year pairs (CVaR_0.50_), ADAPT had higher AUC-CVaR than the comparator methods for nearly all PheCodes within MGB, within Duke, and after MGB-to-Duke transport (Figure 2, left). This advantage was observed broadly across individual mental-health outcomes rather than being driven by a small number of PheCodes (Figure 2, left). For example, within MGB, ADAPT achieved an AUC-CVaR of 0.74 for schizophrenia and other psychotic disorders (PheCode 295), compared with 0.64 for the current-period-only model and 0.69 for maximin. Higher AUC-CVaR was also observed for bipolar disorder (296.1), suicidal ideation (297.1), post-traumatic stress disorder (300.9), and substance addiction and disorders (316). Within Duke, ADAPT achieved an AUC-CVaR of 0.78 for suicide or self-inflicted injury (297.2), compared with 0.58 for the current-period-only model and 0.71 for TransGLM. After MGB-to-Duke transport, ADAPT achieved an AUC-CVaR of 0.71 for schizophrenia and other psychotic disorders, compared with 0.58 for the Duke current-period-only model, corresponding to a 21.5% relative improvement.

This pattern was consistent when different fractions of the lower performance distribution were considered (Figure 2, right). At *α* = 0.20, ADAPT had higher AUC-CVaR_*α*_ than all four comparator methods. The improvement ranged from 0.016 to 0.036 within MGB and from 0.011 to 0.018 within Duke (all *p* < 0.001). Within both MGB and Duke, ADAPT also had higher AUC-CVaR than all four comparators at *α* = 0.40, 0.60, and 0.80 (Supplementary Figures S3-S4). After MGB-to-Duke transport, ADAPT likewise exceeded each transported comparator, with gains ranging from 0.0079 to 0.037, at *α* = 0.2 (all *p* < 0.001). As a contextual benchmark, transported ADAPT also slightly exceeded a current-period-only model trained locally at Duke by 0.0069 (*p* < 0.001).

Threshold-based results similarly suggested that ADAPT improved case detection without sacrificing enrichment among patients identified as high risk. At *α* = 0.20, sensitivity-CVaR for ADAPT exceeded that of the current-period-only model by 4.3, 1.6, and 2.7 percentage points in the within-MGB, within-Duke, and MGB-to-Duke analyses, respectively (all *p* < 0.001). Within MGB, ADAPT also had substantially higher lift-CVaR than the current-period-only, pooled, and maximin models, by 0.23, 0.11, and 0.41, respectively (all *p* < 0.001), while remaining similar to TransGLM. Within Duke, ADAPT improved sensitivity-CVaR over the four comparators by approximately 1.6, 0.9, 0.6, and 0.9 percentage points, respectively (*p* < .001), while also achieving comparable or higher lift-CVaR, with gains of approximately 0.11, 0.065, 0.07, and 0.12. After MGB-to-Duke transport, ADAPT improved sensitivity-CVaR over the transported current-period-only, TransGLM, pooled, and maximin models by approximately 2.7, 2.0, 1.0, and 0.2 percentage points, respectively, and improved lift-CVaR by approximately 0.24, 0.08, 0.28, and 0.19. These patterns were broadly similar at *α* = 0.40, 0.60, and 0.80; the main exceptions were that lift was often comparable between ADAPT and TransGLM, and sensitivity became similar between ADAPT and transported maximin at some larger *α* values (Supplementary Figures S3-S5). Compared with the current-period-only model locally trained at Duke, transported ADAPT had higher lift-CVaR by approximately 0.15 but lower sensitivity-CVaR by 1.47 percentage points at *α* = 0.20.

### 3.3 Case study: suicide-attempt prediction

Following the multi-outcome evaluation, we examined one-year suicide-attempt prediction as a focused case study of model durability (Figure 1C). We evaluated whether ADAPT could maintain contemporaneous discrimination while reducing performance loss as the model aged, both within MGB and after transport to Duke.

**Figure 1:**
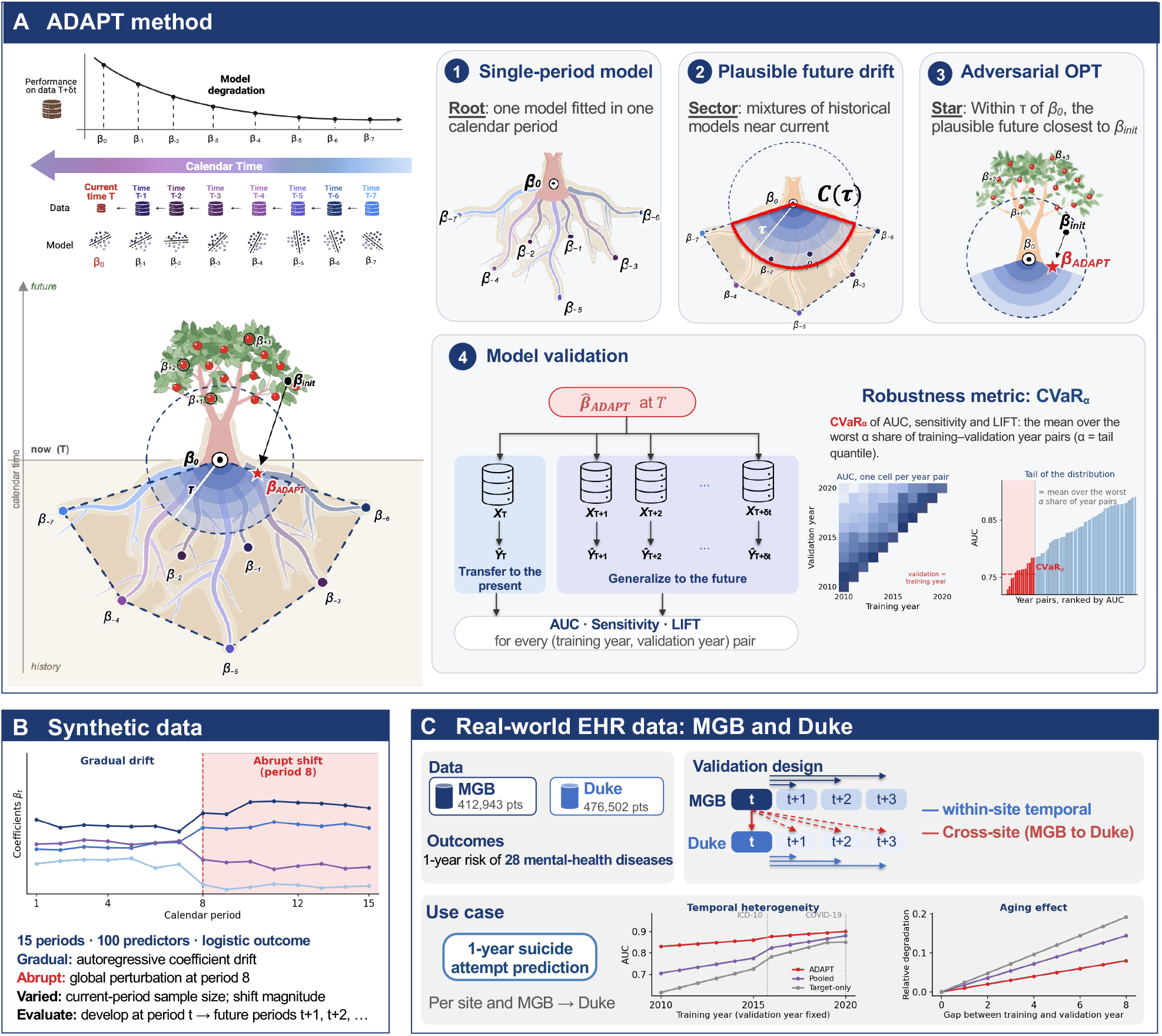
Overview of ADAPT and the study design. (A) ADAPT fits a model in each calendar period (1), defines an uncertainty set *C*(*τ*) of mixtures of current and historical models that remain compatible with held-out current-period data (2), and selects the robust model 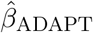 within *C*(*τ*) (3). Models developed at year *T* are evaluated in the current and future years, and performance across training–validation year pairs is summarized by CVaR_*α*_ (4). (B) Synthetic data with gradual model drift and an abrupt shift at period 8. (C) One-year risk of 28 mental health outcomes at MGB and Duke, with within-site temporal and MGB-to-Duke validation.

**Figure 2:**
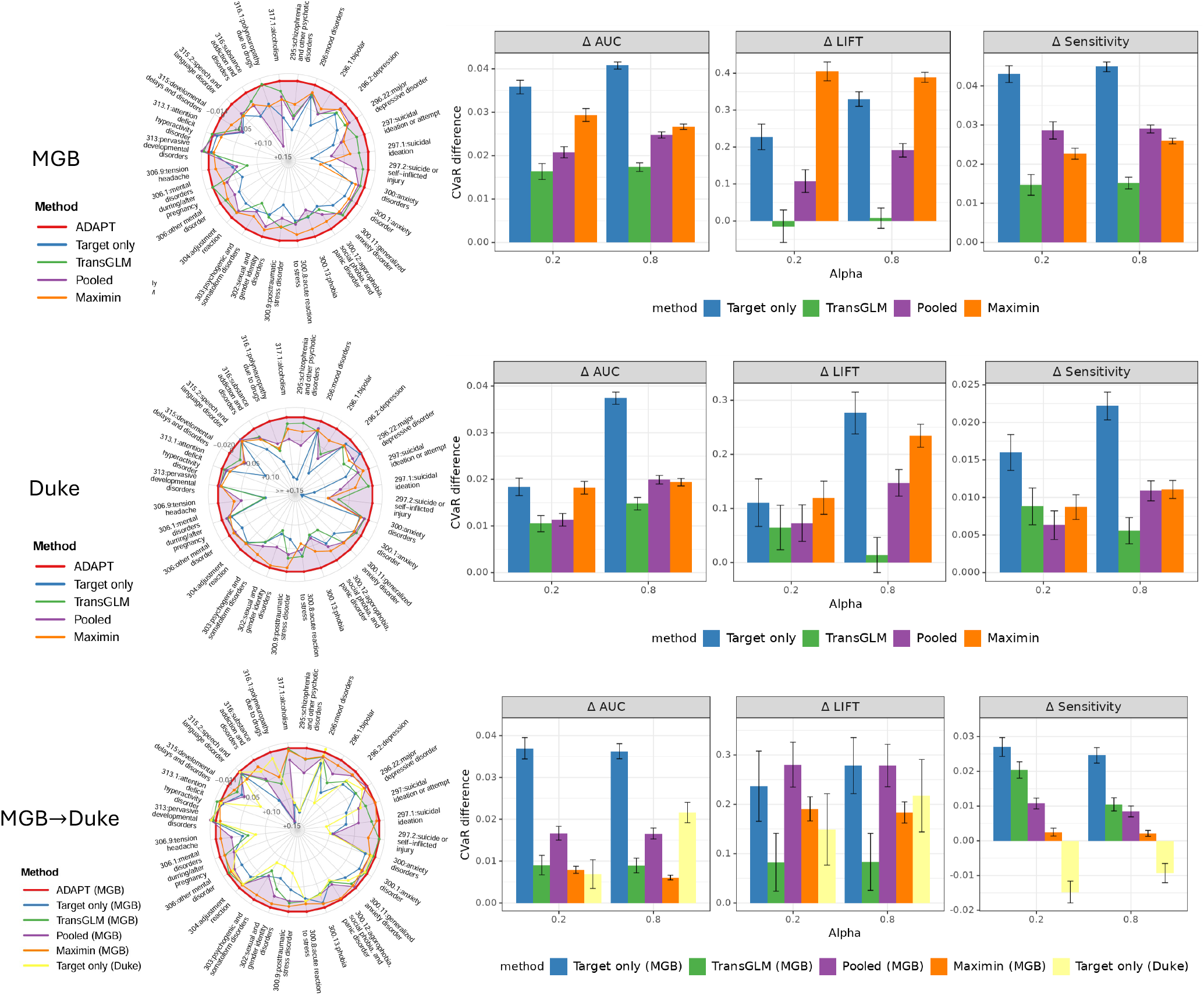
CVaR of benchmarks versus ADAPT within MGB, within Duke, and from MGB to Duke. All methods used the original EHR predictors augmented with year-specific XGBoost prediction scores. Left: radar plots of each phecode’s CVaR of AUC compared to ADAPT at *α* “ 0.5. Right: difference in CVaR (ADAPT - benchmark method) of the phecode-averaged AUC, lift, and sensitivity across *α*. Error bars are 95% confidence intervals, computed as the point estimate ± 1.96 × the standard deviation of the paired bootstrap differences across 500 resamples.

#### 3.3.1 Prediction performance across calendar years and health systems

Because temporal robustness is useful only if strong current-period performance is retained, we first examined contemporaneous discrimination. In contemporaneous internal validation, in which models were developed and evaluated within the same MGB calendar year, ADAPT achieved the highest mean AUC (0.815; Figure 3, left). The cumulative pooled-data model had a mean AUC of 0.800, followed by TransGLM at 0.794, the current-period-only model at 0.763, and maximin at 0.730. The advantage of ADAPT was most apparent in the earlier target years, when the current-period data contributed less information than the accumulated historical periods. This advantage was retained when the MGB-developed models were transported directly to Duke patients from the corresponding calendar year: mean AUC was 0.85 for ADAPT, compared with 0.82 for TransGLM, 0.82 for the pooled-data model, 0.70 for the current-period-only model, and 0.75 for maximin (Figure 3, right). Thus, the robustness incorporated by ADAPT did not come at the expense of contemporaneous discrimination and remained beneficial after cross-health-system transport.

**Figure 3:**
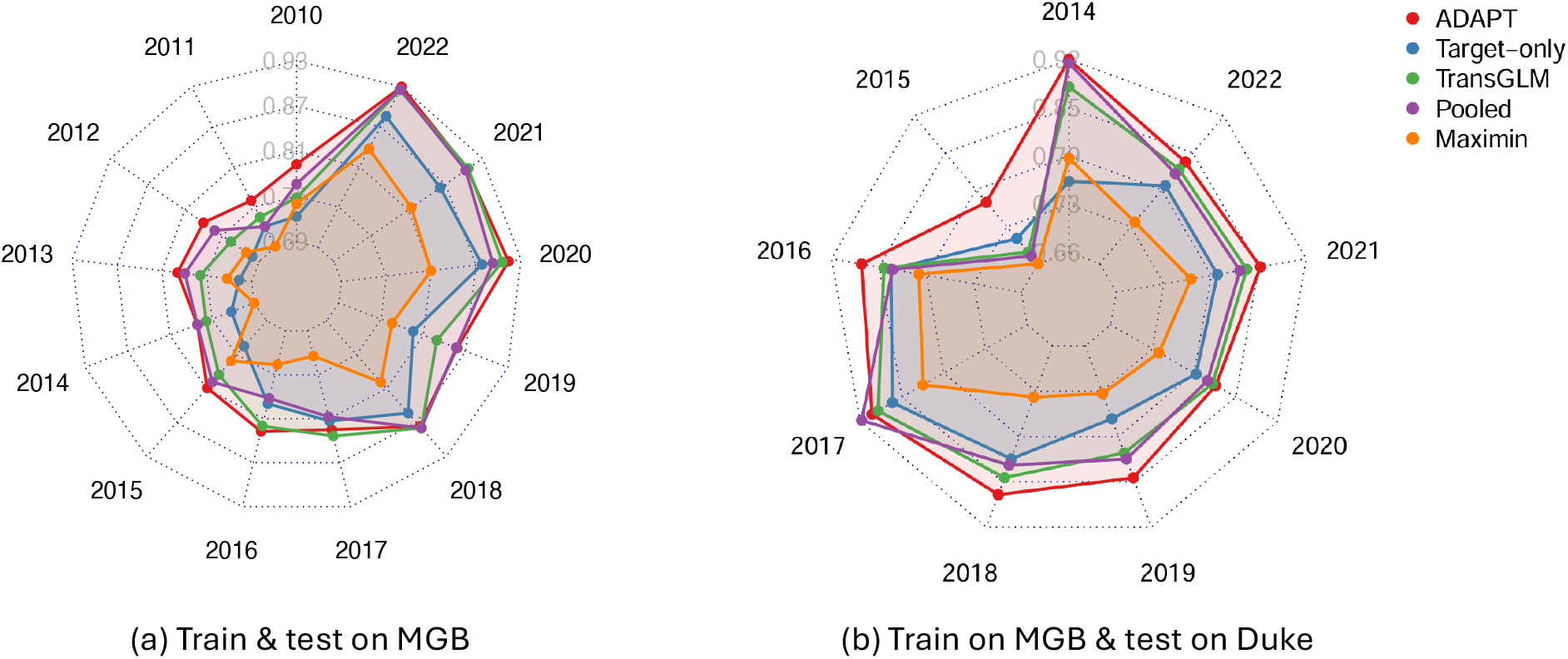
Contemporaneous discrimination. Models were developed for a target calendar year at MGB and evaluated either in MGB data from the same calendar year (left) or in Duke data from that calendar year (right). Points show mean AUC across repeated splits.

We next examined the effect of model age more directly by holding the validation population fixed and using models developed in progressively earlier years. This separated the effect of older development data from changes in the validation population itself. In a fixed 2020 validation population (Figure 4), ADAPT maintained the highest mean AUC across development years at both MGB (0.881 [SD over different training-year models: 0.019]) and Duke (0.800 [0.013]). The best-performing comparator was TransGLM at MGB (0.850 [0.031]) and the pooled-data model at Duke (0.771 [0.026]). Although maximin varied less across development years, its mean AUC remained lower at both sites: 0.808 at MGB and 0.730 at Duke. This contrast illustrated that stability across model ages alone was insufficient if achieved at a substantially lower level of discrimination.

**Figure 4:**
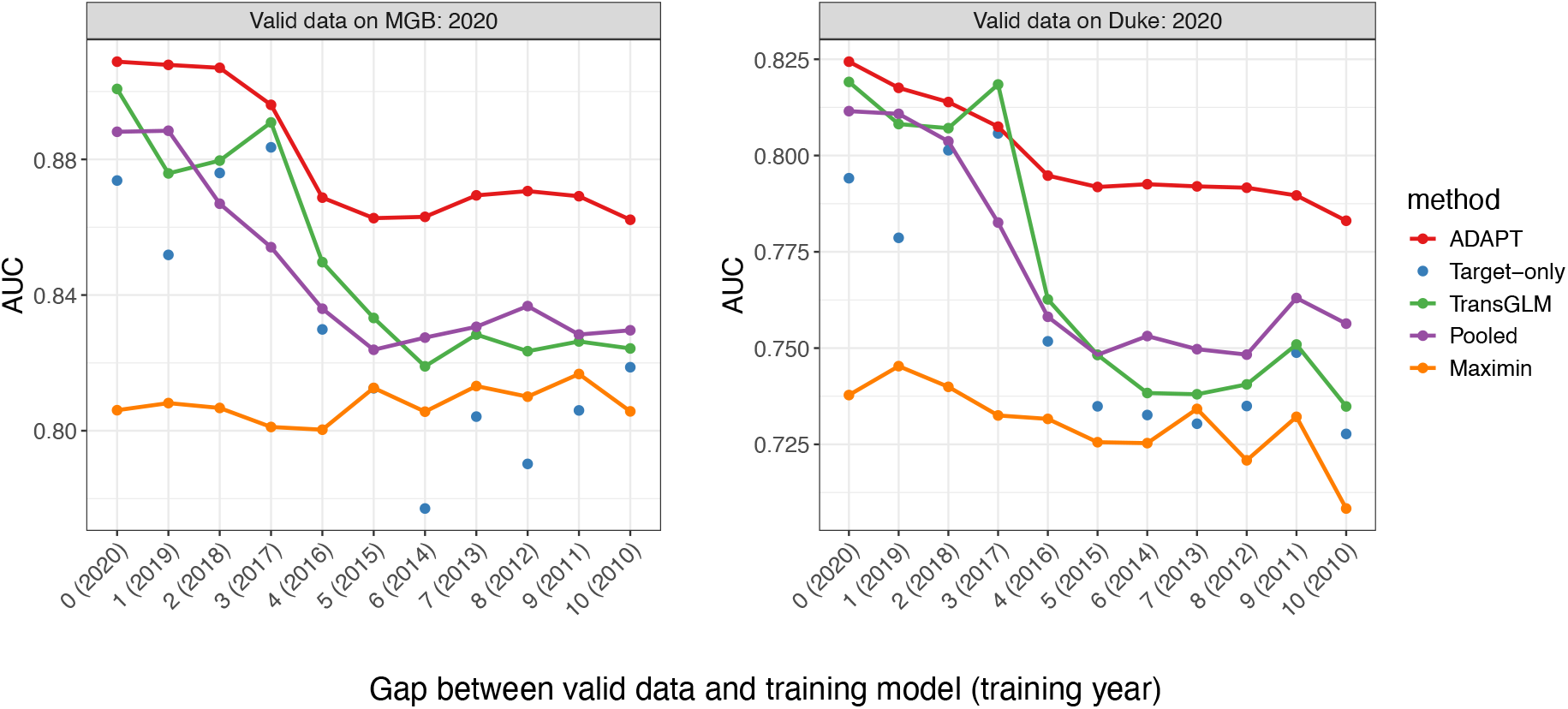
Temporal validation in a common 2020 evaluation population. Models were developed for progressively earlier target years and evaluated in MGB 2020 data (left) or Duke 2020 data (right).

#### 3.3.2 Temporal durability

We next assessed whether ADAPT lost less accuracy as the model aged. Each older model was compared with its corresponding “newborn” model, developed using data from the validation year itself. Both models were evaluated in the same held-out population, so that the comparison reflected the effect of model age. For external validation, both models were developed at MGB and evaluated in Duke data from the newborn model’s training year.

At MGB, ADAPT’s relative temporal degradation was 2.57% after two years and 7.65% after eight years, compared with 5.14% and 12.1%, respectively, for the pooled-data model (Figure 5, left). Both the current-period-only model and TransGLM exceeded 17% degradation after eight years. Maximin showed less long-term degradation than ADAPT—5.16% after eight years—but from a substantially lower baseline: the lowest AUC among the corresponding newborn models was 0.721 for maximin and 0.833 for ADAPT. Thus, maximin’s smaller relative decline reflected stability from a substantially lower starting level of discrimination.

**Figure 5:**
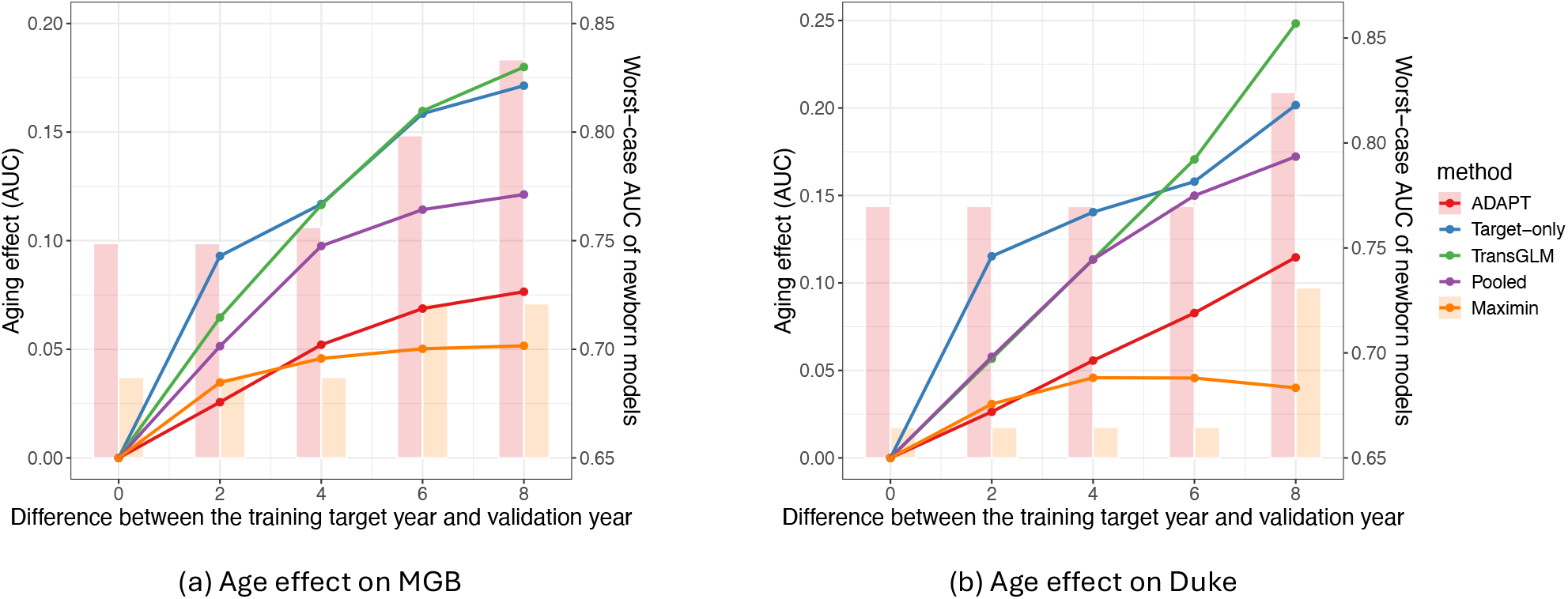
Relative temporal degradation as the interval between model development and evaluation increased. Left: temporal validation at MGB. Right: MGB-developed models evaluated at Duke. Bars show the worst (“newborn”) AUC within each interval window for ADAPT and maximin, illustrating that low relative degradation can coexist with low baseline discrimination.

The same trade-off between discrimination and temporal stability was observed after transport to Duke (Figure 5, right). ADAPT’s relative degradation after two years was 2.64%, compared with 5.67% for TransGLM and 11.5% for the current-period-only model. TransGLM’s degradation increased to 24.8% after eight years. Maximin again showed smaller long-term degradation but had the lowest worst-case newborn AUC. Together, these findings indicated that ADAPT retained its balance of strong discrimination and reduced temporal deterioration after cross-health-system transport.

As a sensitivity analysis, we examined whether the relative performance patterns across methods were preserved when the original EHR predictors were augmented with nonlinear features from year-specific XGBoost models. ADAPT remained among the best-performing methods for contemporaneous discrimination and continued to show less temporal degradation than the current-period-only and pooled-data approaches under the augmented feature sets (Supplementary Figures S6–S8). These findings suggested that the relative advantage of ADAPT was not specific to the original linear feature representation.

### 3.4 Synthetic data results

In the synthetic-data study, ADAPT performed well across current-period, future-generalization, and abrupt-shift settings. When models were evaluated in the same period used as the training target, ADAPT matched TransGLM with very limited current-period data (*ρ* = 0.1) and outperformed the other comparators; as *ρ* increased, its performance approached that of the target-only model (Figure S1, left). When models were instead evaluated on subsequent periods, ADAPT consistently achieved the highest worst future-period AUC across all values of *ρ*, while TransGLM was competitive mainly when current-period data were limited and pooling and maximin remained less accurate (Supplementary Figure S1, right). Under the abrupt perturbation introduced at period 8 to represent a regime change, performance was generally higher when model development and validation occurred within the same regime (pre–pre or post–post) than when they spanned the shift (Supplementary Figure S2). The largest degradation occurred when models developed before the perturbation were evaluated after it, and worsened as *p*_perturb_ increased. ADAPT deteriorated more slowly and maintained the highest median AUC in this setting. Pooling and TransGLM performed competitively when both development and validation were pre-shift but declined more sharply across the pre-to-post shift; the target-only model generalized poorly to post-shift data, whereas maximin remained comparatively stable but at a consistently lower level of discrimination.

## 4 Discussion

In this retrospective multi-health-system study, ADAPT was developed to address a practical problem in EHR-based mental health risk prediction: a model that performs well for patients seen at development can deteriorate as clinical populations and data-generating processes change. ADAPT combined historical period-specific models with limited current-period data and explicitly balanced current accuracy against adverse performance under plausible future relationships. Across 28 mental-health outcomes, ADAPT generally achieved stronger performance in the more challenging temporal settings within both health systems and after cross-health-system transport. In the focused suicide-attempt case study, ADAPT maintained strong contemporaneous discrimination while showing less performance degradation as the interval between model development and evaluation increased. Conventional maximin-based DRO learning was sometimes more stable over long intervals, but its lower baseline AUC showed that stability alone does not ensure clinically useful prediction.

The findings highlight a distinction between adapting to the present and preparing for the near future. A current-period-only model directly represents recent patients but can be imprecise when the contemporary sample contains few outcomes. Cumulative pooling increases sample size but treats older and recent data as though they were exchangeable. TransGLM borrows from historical periods to improve a specified current target, but it does not explicitly optimize durability after that target period. ADAPT uses held-out current-period data to exclude historical mixtures that no longer fit the present and then protects against the most adverse model within the remaining range. The multi-outcome analyses showed that this strategy improved performance across a broad range of mental-health prediction tasks. The suicide-attempt case study further showed that the same approach could preserve contemporaneous discrimination while reducing deterioration as a model aged. Its advantage was also observed when models developed at MGB were evaluated at Duke, despite differences in patient composition and documentation environments.

Temporal robustness is especially relevant for suicide-attempt prediction. Recorded diagnoses, medication use, health-care encounters, and suicide-related codes reflect both underlying patient risk and the clinical processes through which that risk is observed. Changes in screening, coding, treatment, access to care, and help-seeking can therefore affect both predictor distributions and their associations with documented outcomes. ADAPT does not identify the cause of an observed shift, nor does it remove the need for model monitoring. Instead, it uses heterogeneity already observed across calendar periods to reduce dependence on a single development distribution. The summary-level implementation also means that historical periods or institutions need not contribute raw patient-level data after their models have been estimated, although labeled current-period data are still required.

The sensitivity analysis using XGBoost-derived nonlinear features produced similar relative performance patterns across methods, suggesting that ADAPT’s advantage was not specific to the original feature representation. More generally, because ADAPT operates on a flexible feature map, the framework can be applied to representations derived from more complex prediction models while retaining the same strategy for learning from temporal heterogeneity. The synthetic-data experiments provided complementary evidence under controlled shifts: ADAPT maintained current-period performance when contemporary data were limited, achieved the highest worst future-period AUC across varying amounts of current data, and deteriorated more slowly when models developed before an abrupt regime change were evaluated afterward. These results help separate the intended robustness of the method from characteristics specific to the observed EHR cohorts.

The study has several strengths. It evaluated model durability over many calendar years rather than reporting only a random train–test split. It assessed performance across 28 mental-health outcomes and then examined temporal degradation in greater depth in a suicide-attempt case study. The study period spanned major changes in coding and care delivery, and an independent Duke cohort provided external validation. Comparisons included recent-period training, cumulative pooling, transfer learning, and conventional worst-case learning, allowing current accuracy and temporal stability to be considered jointly. Controlled simulations complemented the EHR analyses by separating gradual coefficient evolution from an abrupt perturbation.

Several limitations should be considered. First, this was a retrospective analysis of EHR-documented outcomes. Events occurring outside MGB or Duke might not have been captured, and temporal changes in documentation, screening, or care-seeking could appear as changes in outcome risk. Second, although the multi-outcome analysis included discrimination and threshold-based measures, these retrospective metrics do not establish clinical utility, calibration at specific deployment thresholds, or net benefit in practice. Third, external validation involved one additional health system; performance in other regions, care settings, and EHR environments remains unknown. Fourth, ADAPT defines plausible future conditional models through mixtures of historical and current models and therefore can protect only against changes represented by, or sufficiently related to, the historical variation observed during development. Unprecedented shifts outside this range can still lead to performance loss. Finally, this study did not assess prospective workflow integration, clinician response, patient acceptability, fairness across demographic groups, or effects on patient outcomes.

ADAPT therefore should be viewed as a component of model stewardship rather than a substitute for surveillance or clinical judgement. Before deployment, a health system would still need to select clinically meaningful thresholds, evaluate calibration in the intended population, monitor performance and data quality, and define conditions for recalibration or retraining. Prospective evaluation is needed to determine whether improved temporal durability translates into more reliable clinical assessment and better patient outcomes. Within these limits, our findings show that temporal heterogeneity already present in historical EHR can be used to improve the durability of mental-health risk prediction while preserving performance for contemporary patients.

## Data Availability

All data produced in the present study are available upon reasonable request to the authors.

## Data sharing

The EHR data from MGB and Duke that support the findings of this study are available from the respective health systems, but restrictions apply because the data were used under license and are not publicly available. Access may be available upon request and with permission from the relevant institutions.

## Funding

This research did not receive funding. The role of the funding source was therefore not applicable.

## Declaration of interests

The authors declare no competing interests or other interests that might be perceived to influence the results or discussion reported in this paper.

## Supplementary material

### A Supplementary methods

#### A.1 Formal setup and notation

Let *Y* denote a binary, continuous, or count outcome and let *X* denote a vector of observed predictors or features derived from a pretrained model. ADAPT assumes access to labeled data from *L* historical source populations,

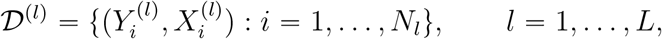

and a labeled current target dataset

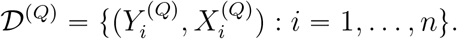

For source *l*, the conditional outcome model is indexed by ***β***^(*l*)^ and has mean

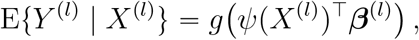

where *g* is a known link and *ψ*(·) is a prespecified feature map. The feature map can be the original predictor vector, a pretrained risk score, or a representation such as the last-layer features of a pretrained neural network. The current target conditional model is indexed by ***β***^(*Q*)^. A future target is assumed to have the same model form but may have a shifted coefficient vector ***β***^(*T*)^ ≠ ***β***^(*Q*)^. Let 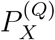 denote the current target covariate distribution.

#### A.2 ADAPT algorithm

ADAPT has three steps: estimation of historical and current-period models; construction of an uncertainty set for plausible future models; and adversarial optimization using a second-order approximation. Only model-level summaries are required from the historical sources. The current target dataset is randomly divided into two disjoint, equally sized subsets, 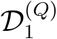 and 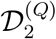. The first subset is used to estimate the current-period model, and the second is used to evaluate candidate future models. Splitting the current target data prevents the same observations from being used both to estimate the target anchor and to determine which mixtures are compatible with the target.

**Step 1: estimation of historical and current-period models**. For each source *l*, estimate

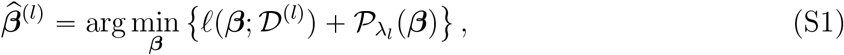

where *ℓ* is the negative log-likelihood and 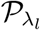 is a user-specified penalty, such as the elastic net, with tuning selected by standard procedures including cross-validation. Estimate the current target model from 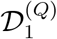 by

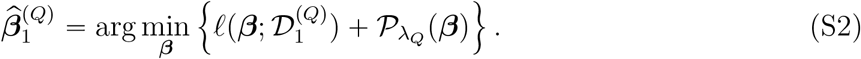

**Step 2: uncertainty set of plausible future models**. Define

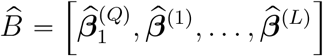

and let

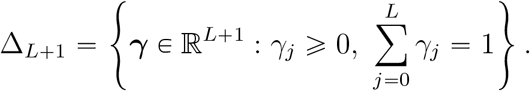

Candidate future parameters have the form 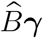. Candidates are retained when their loss on the held-out current target subset does not exceed *τ*:

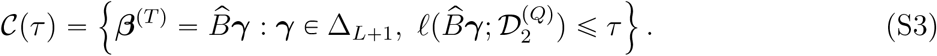

A smaller *τ* restricts the uncertainty set to mixtures that closely match the current target data; a larger *τ* allows broader departure from the current target.

Let 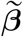 be the convex combination of historical source coefficients that minimizes loss on 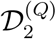.

The threshold used in the study was

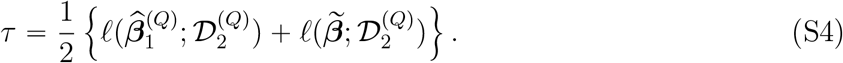

This choice uses both the held-out performance of the current-period model and the best historical-source combination to determine the breadth of the uncertainty set. A high held-out loss for the current-period model can reflect imprecision from limited current data, whereas a high loss for the source combination can reflect substantial departure of the current target from historical periods. In either case, the construction avoids relying only on a potentially noisy current-period estimator.

**Step 3: adversarial optimization**. ADAPT selects a model by minimizing worst-case excess loss over *C*(*τ*):

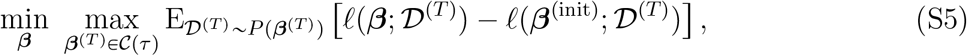

where

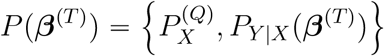

and ***β***^(init)^ is a reference estimator. In the analyses, the reference was taken as the sample-size-weighted average of the period-specific estimates. The objective therefore evaluates performance relative to the reference model under adverse but plausible future conditional relationships while retaining the current target covariate distribution.

Direct optimization is simplified by a second-order Taylor approximation around ***β***^(init)^:

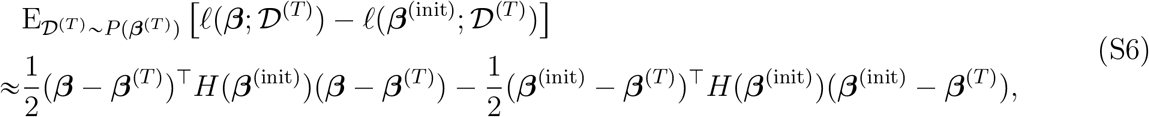

where *H*(***β***^(init)^) is the Hessian of the negative log-likelihood evaluated at ***β***^(init)^. Since *C*(*τ*) is convex, the minimax problem under this quadratic approximation is equivalent to projecting ***β***^(init)^ onto *C*(*τ*) under the geometry induced by *H*(***β***^(init)^). The resulting estimator is

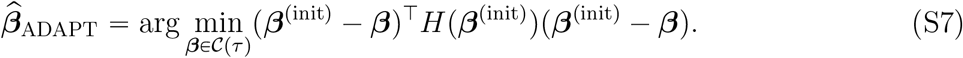

Thus, the one-step estimator can be interpreted as a projection of the initial reference model onto the restricted set of plausible future models under the geometry induced by the likelihood Hessian.

#### A.3 XGBoost nonlinear feature set

The nonlinear augmentation analysis used fitted XGBoost prediction scores as additional predictors in the regression models. Rather than averaging the historical XGBoost predictions into one score, ADAPT retained a separate score from each historical year and appended these scores to the original clinical predictors. This allowed the downstream regression to use nonlinear patterns learned in different calendar periods while retaining the ADAPT procedure for combining period-specific regression models.

Within each repetition, data from each calendar year *l* were divided into three disjoint subsets,

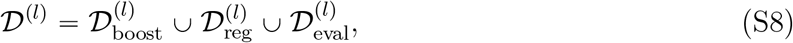

with one quarter used to train XGBoost models, one quarter used for the regression-based methods, and one half held out for evaluation. XGBoost models were fitted only on the boosting subsets and applied to the regression and evaluation subsets. Thus, a prediction score used for regression fitting or evaluation was generated by an XGBoost model that had not been trained on that observation. The inverse probability of censoring weights described in the main Methods were used in model fitting.

Let *X* ∈ ℝ^*p*^ denote the original predictor vector, *A*_XGB_ denote the XGBoost fitting procedure. The year-specific XGBoost prediction score trained on 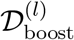 can be defined as

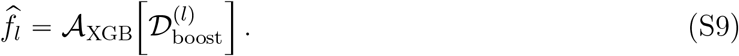

Applying every historical XGBoost model trained at or before year *l* to the same predictor vector *x* gives the score vector, which was combined to form the augmented feature map *ψ*_*l*_(*x*):

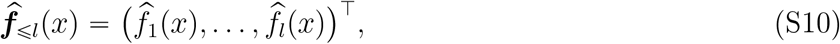

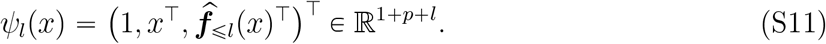

Note that different benchmarks used different components of 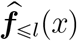 given their assumed data access. The current-period-only model had access only to current-year data and therefore appended the single score 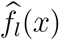 from the XGBoost model trained in year *l*:

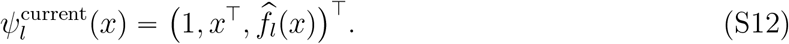

In contrast, ADAPT, maximin, and transGLM used historical-source information and were given exactly the same additional predictors: the full vector of year-specific historical scores 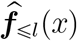 defined in Equation (S10). Their feature maps were therefore

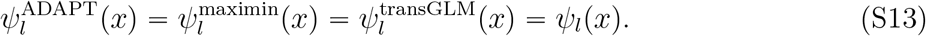

All three retained each historical score as a separate predictor. Thus, their differences arose from downstream regression fitting and model combination, rather than different XGBoost feature sets. The pooled-data model assumed access to observations that could be combined across its eligible years. Therefore, we appended one score from the XGBoost model trained on those pooled observations:

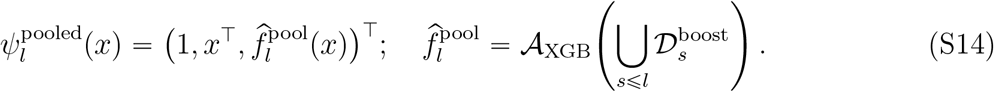

For temporal validation in a later year *v* ≥ *l*, each method retained its own development-year feature map, fitted score-generating models, and regression coefficients. The current-period-only model therefore continued to use 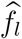, not a newly fitted 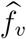, and the historical score library for ADAPT, maximin, and TransGLM remained fixed at 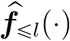. The pooled score model likewise remained unchanged. This comparison measures aging of the complete prediction pipeline, including its nonlinear feature representation.

#### A.4 Synthetic Data Generation and Results

##### A.4.1 Design for Synthetic Data Generation

At discrete periods *l* ∈ {1, … , *L*} with *L* = 15, independent datasets {*X*^(*l*)^, *Y* ^(*l*)^} of *N* = 2000 observations were generated. Each predictor vector followed 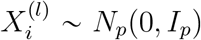 with *p* = 100, and the binary outcome followed 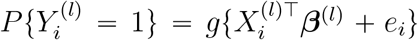, where *g* is the logistic function and *e*_*i*_ ∼ *N* (0, 1) is subject-level noise. An independent validation dataset of the same size was generated in each period from the same coefficient sequence.

The coefficient sequence combined gradual autoregressive evolution of order *m* = 4 with coordinate-specific stochastic shocks. The initial *m* coefficient vectors were drawn from *N*_*p*_(0, *I*_*p*_), with *p*_0_ = 30 randomly selected coordinates set to zero. For later periods,

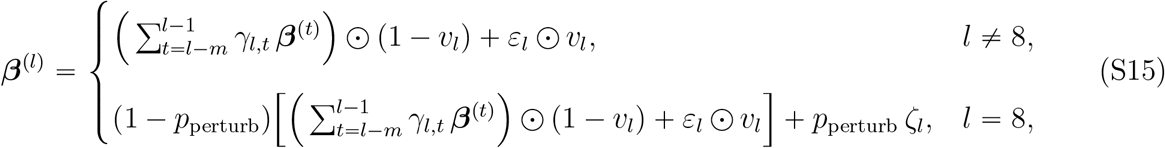

where ⊙ denotes element-wise multiplication. For each period, the weights (*γ*_*l,l*−*m*_, … , *γ*_*l,l*−1_) were drawn independently from a Dirichlet(1, 1, 1, 1) distribution. Each element of *v*_*l*_ was drawn independently from Bernoulli(*p*_shock_) with *p*_shock_ = 0.3, *ε*_*l*_ ∼ *N*_*p*_(0, *I*_*p*_), and *ζ*_*l*_ ∼ *N*_*p*_(0, 0.5*I*_*p*_). Thus, in each period about 30% of coefficients were replaced by independent draws, while the remainder evolved as convex combinations of the preceding four periods. The process was run for a burn-in of 10 periods before the final *L* = 15 periods were retained, so that results did not depend on the initial values. A single coefficient trajectory was generated and held fixed across Monte Carlo repetitions. The perturbation at period 8 represented an abrupt regime change.

Each period *t* ∈ {6, … , 15} served in turn as the current target period, with all earlier periods 1, … , *t* − 1 as historical sources. In order to mimic the scenario with scarce target data, within the target period, a random subsample of size *ρN* was selected to be the training target data. To assess the effect of the amount of current-period data on model performance, *ρ* was varied over {0.1, 0.2, 0.3, 0.5, 1} with *p*_perturb_ = 0. To assess resilience to the regime change, *p*_perturb_ was varied from 0 to 0.9 with *ρ* = 0.2. Period-specific models were fitted by *l*_1_-penalised logistic regression, with the penalty selected by 10-fold cross-validation using glmnet. The pooled-data model applied the same procedure to all observations from periods 1, … , *t* − 1 together with the current-period training subsample. TransGLM was fitted with the `glmtrans` package using its default tuning and source detection. The maximin estimator solved 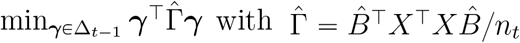, where 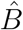 collects the source coefficient estimates.

A model developed at period *t* was evaluated on the validation data from every period *v* ∈ {*t*, … , 15}. Current-period performance was the AUC at *v* = *t*, and future-period performance was summarized by the minimum AUC across validation periods for each development period. For the perturbation analysis, development and validation periods were grouped as before (*l* ⩽ 7) or after (*l* ⩾ 8) the perturbation. All AUCs were averaged over 50 Monte Carlo repetitions. Figure S1 reports the mean across development periods, and each boxplot in Figure S2 shows the distribution of AUC across the training–validation period pairs within the corresponding pre- or post-perturbation group.

##### A.4.2 Detailed Results from Synthetic Data Study

###### Performance on the current target population

When validation data come from the same period as the training target (Figure S1, left), ADAPT consistently benefits from historical sources. Even with limited target data, i.e., ratio of the current data sample size to the historical sample size *ρ* = 0.1, the performance of ADAPT matches transGLM and exceeds other baselines. As *ρ* increases, ADAPT converges to the target-only estimator, while pooling and maximin remain insensitive to *ρ* and consistently underperform. TransGLM performs comparably to ADAPT when *ρ* = 0.1, but falls below target-only once *ρ* ⩾ 0.3.

**Figure S1:**
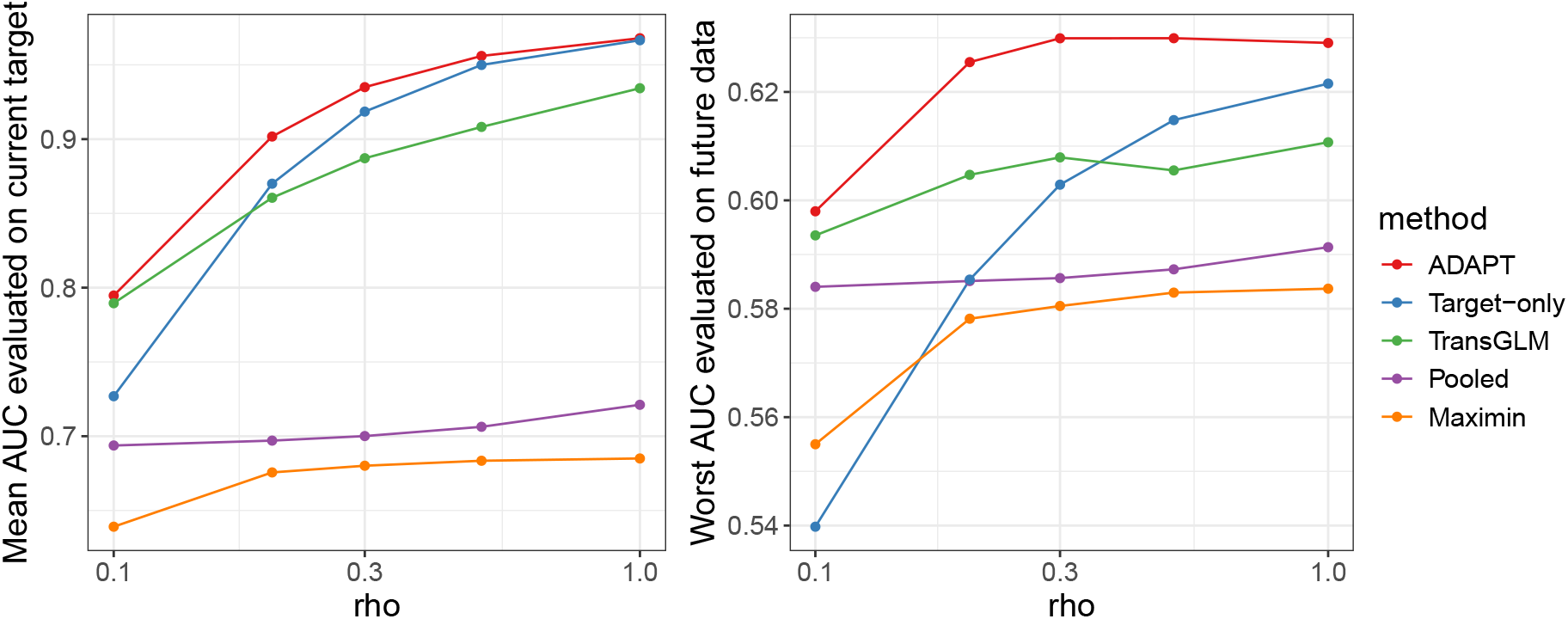
AUC as the current-to-historical data ratio increases. Left: validation data come from the same period as the training data; right: validation data come from periods after the training data.

###### Generalization to future populations

To assess robustness under temporal shift, we applied models trained at period *t* to data from subsequent periods *t*^′^ > *t* and summarized performance by the worst AUC achieved across future periods (Figure S1, right). This captures how well models generalize once deployed on unseen data subject to distributional changes. ADAPT consistently attains the highest worst-case AUC on future data sets for all *ρ*, indicating greater stability under temporal shift. Maximin only surpasses target-only when *ρ* = 0.1 but otherwise yields the lowest generalizability, while transGLM ranks second for *ρ* ⩽ 0.3 before falling below target-only. Pooling remains relatively stable but consistently below ADAPT and transGLM.

###### Generalization under perturbed design

We further assessed robustness by partitioning training and validation into pre-shift (*t* ⩽ 7) and post-shift (*t* ⩾ 8) periods (Figure S2). Evaluations within the same period type (pre-pre or post-post) yield higher AUC than those spanning the shift. Training on pre-shift data and validating on post-shift data shows the greatest degradation, which worsens with increasing *p*_perturb_, reflecting the difficulty of anticipating future changes. ADAPT deteriorates more slowly in this setting and maintains the highest medians, indicating robustness to post-shift distributions. Within-regime evaluations are less sensitive to *p*_perturb_, though pooling and maximin continue to decline even when trained and validated post-shift, suggesting limited forward generalizability. Pooling and transGLM perform competitively when both training and validation are pre-shift, but drop sharply when applied across pre-to post-shift periods. The target-only model shows consistently poor generalizability to post-shift data, while maximin remains uniformly low across all settings, reflecting over-conservativeness.

**Figure S2:**
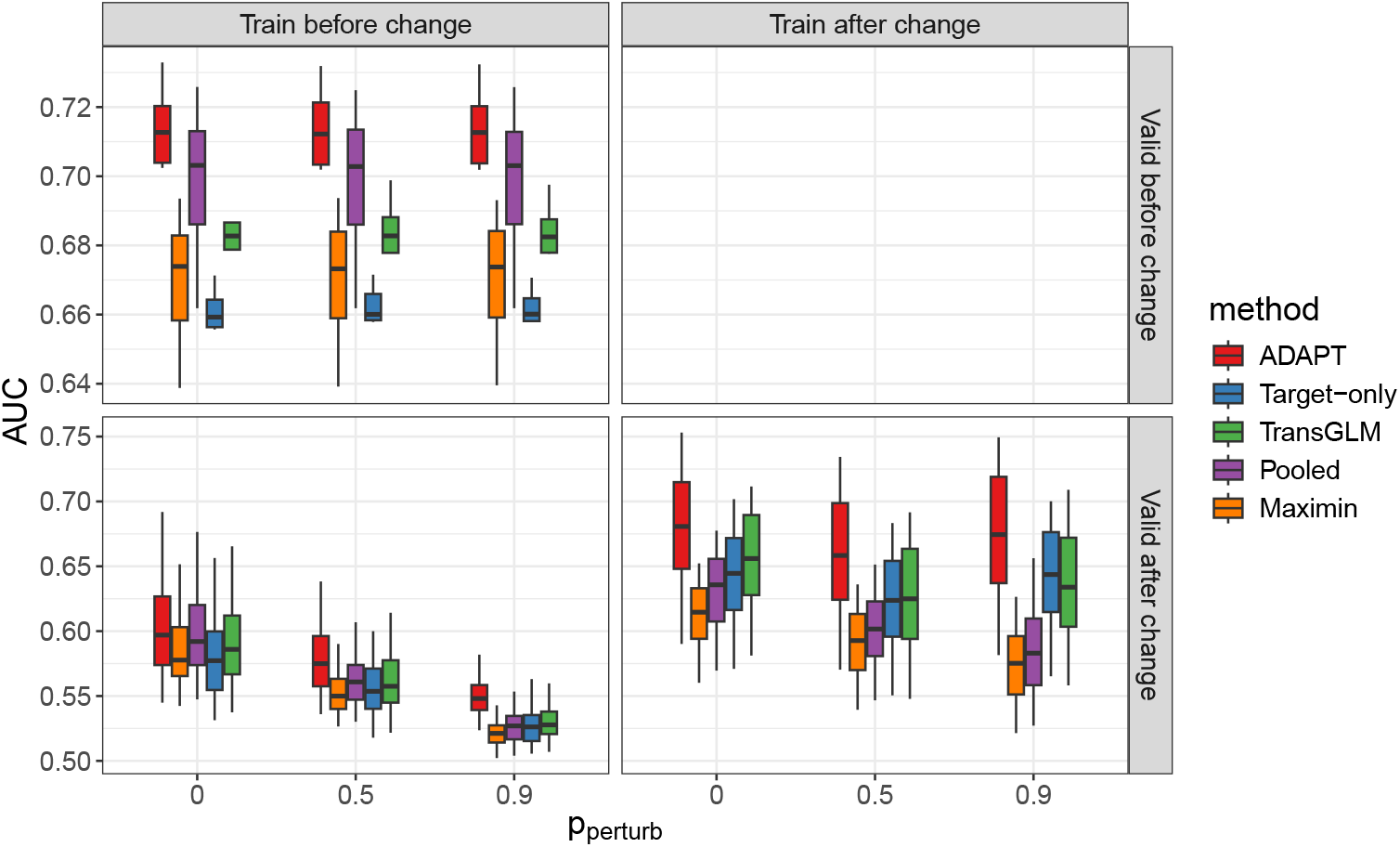
AUC as perturbation level increases. Time window for the training target and validation data is split into two groups: before perturbation (*t* ⩽ 7) and after perturbation (*t* ⩾ 8)

#### A.5 Clinical cohort characteristics and additional analyses

**Table S1:** Baseline characteristics of the Duke and MGB cohorts. Values are *n* (%).

| | Duke ($N = 476,502$) | MGB ($N = 412,943$) | Overall ($N = 889,445$) |
| --- | --- | --- | --- |
| **Gender** |  |  |  |
| Female | 269477 (56.6%) | 266318 (64.5%) | 535795 (60.2%) |
| Male | 207020 (43.4%) | 146625 (35.5%) | 353645 (39.8%) |
| Unknown | 5 (0.0%) | 0 (0%) | 5 (0.0%) |
| **Race** |  |  |  |
| Asian | 10990 (2.3%) | 11008 (2.7%) | 21998 (2.5%) |
| Black | 129915 (27.3%) | 24682 (6.0%) | 154597 (17.4%) |
| White | 305195 (64.0%) | 328755 (79.6%) | 633950 (71.3%) |
| Other | 17318 (3.6%) | 47367 (11.5%) | 64685 (7.3%) |
| Unknown | 13084 (2.7%) | 1131 (0.3%) | 14215 (1.6%) |
| **Age group** |  |  |  |
| 0–18 | 73749 (15.5%) | 27872 (6.7%) | 101621 (11.4%) |
| 19–25 | 34920 (7.3%) | 35919 (8.7%) | 70839 (8.0%) |
| 26–55 | 187999 (39.5%) | 193528 (46.9%) | 381527 (42.9%) |
| > 55 | 179834 (37.7%) | 155624 (37.7%) | 335458 (37.7%) |
| **Landmark year** |  |  |  |
| 2005 | 0 (0%) | 10029 (2.4%) | 10029 (1.1%) |
| 2006 | 0 (0%) | 10859 (2.6%) | 10859 (1.2%) |
| 2007 | 0 (0%) | 11259 (2.7%) | 11259 (1.3%) |
| 2008 | 0 (0%) | 11980 (2.9%) | 11980 (1.3%) |
| 2009 | 0 (0%) | 13355 (3.2%) | 13355 (1.5%) |
| 2010 | 0 (0%) | 14380 (3.5%) | 14380 (1.6%) |
| 2011 | 0 (0%) | 15355 (3.7%) | 15355 (1.7%) |
| 2012 | 0 (0%) | 17128 (4.1%) | 17128 (1.9%) |
| 2013 | 0 (0%) | 17854 (4.3%) | 17854 (2.0%) |
| 2014 | 62705 (13.2%) | 20125 (4.9%) | 82830 (9.3%) |
| 2015 | 79191 (16.6%) | 21335 (5.2%) | 100526 (11.3%) |
| 2016 | 65349 (13.7%) | 25714 (6.2%) | 91063 (10.2%) |
| 2017 | 59416 (12.5%) | 33165 (8.0%) | 92581 (10.4%) |
| 2018 | 52747 (11.1%) | 36727 (8.9%) | 89474 (10.1%) |
| 2019 | 51584 (10.8%) | 40224 (9.7%) | 91808 (10.3%) |
| 2020 | 40649 (8.5%) | 38474 (9.3%) | 79123 (8.9%) |
| 2021 | 38324 (8.0%) | 42218 (10.2%) | 80542 (9.1%) |
| 2022 | 26537 (5.6%) | 32762 (7.9%) | 59299 (6.7%) |

### B Supplementary figures

**Figure S3:**
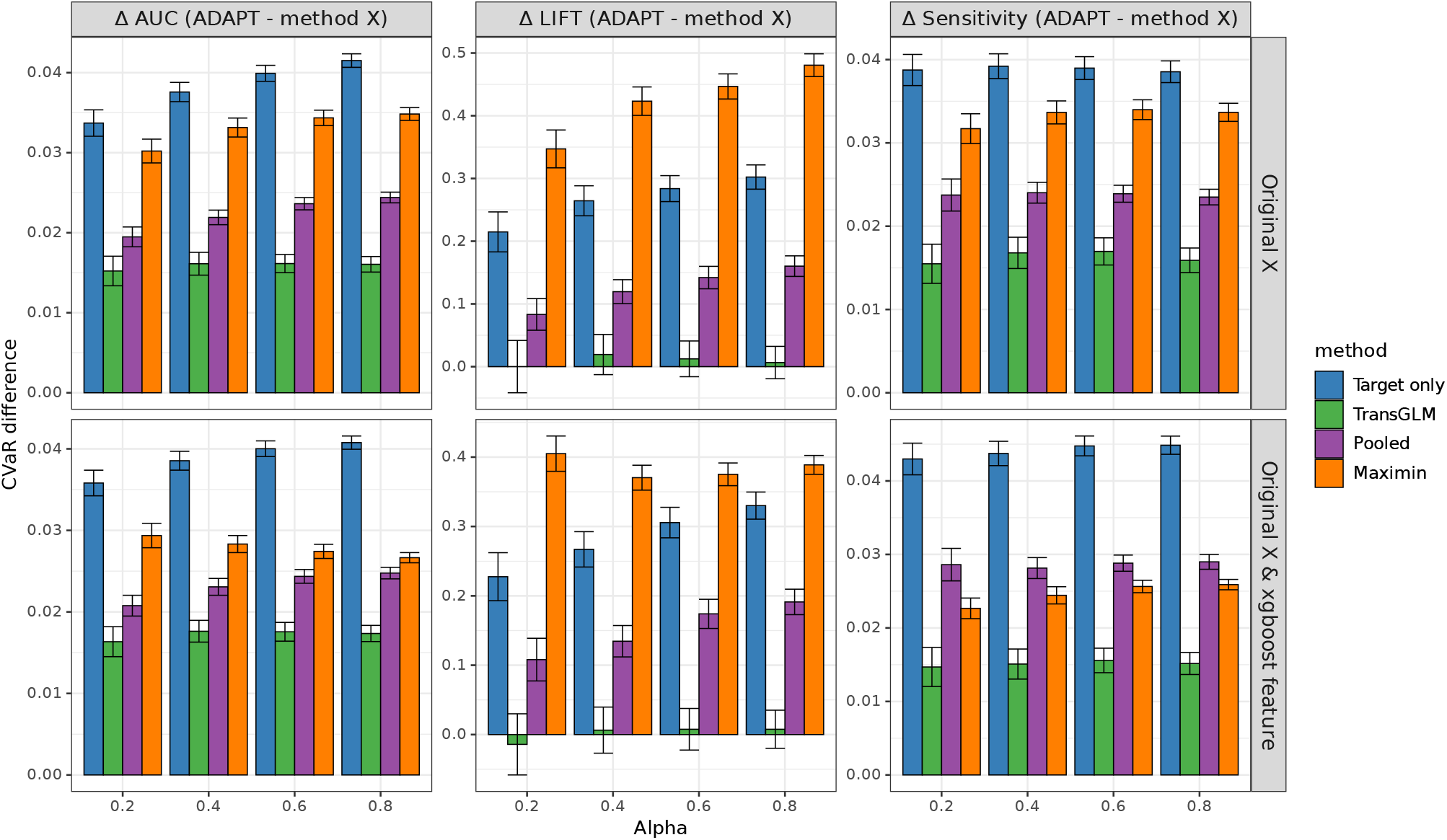
Sensitivity analysis: effect of nonlinear feature augmentation within MGB. For each benchmark method, bars show the difference in lower-tail CVaR between the benchmark and ADAPT 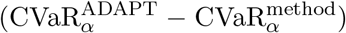, averaged across mental health outcomes, at tail fractions *α* = 0.2, 0.4, 0.6, 0.8, for AUC, lift, and sensitivity in within-MGB temporal validation. Positive values indicate worse worst-case performance than ADAPT. Methods in the top panel used the original predictors, while the lower panel additionally included the XGBoost-derived prediction scores. Error bars are 95% confidence intervals from 500 paired bootstrap replicates.

**Figure S4:**
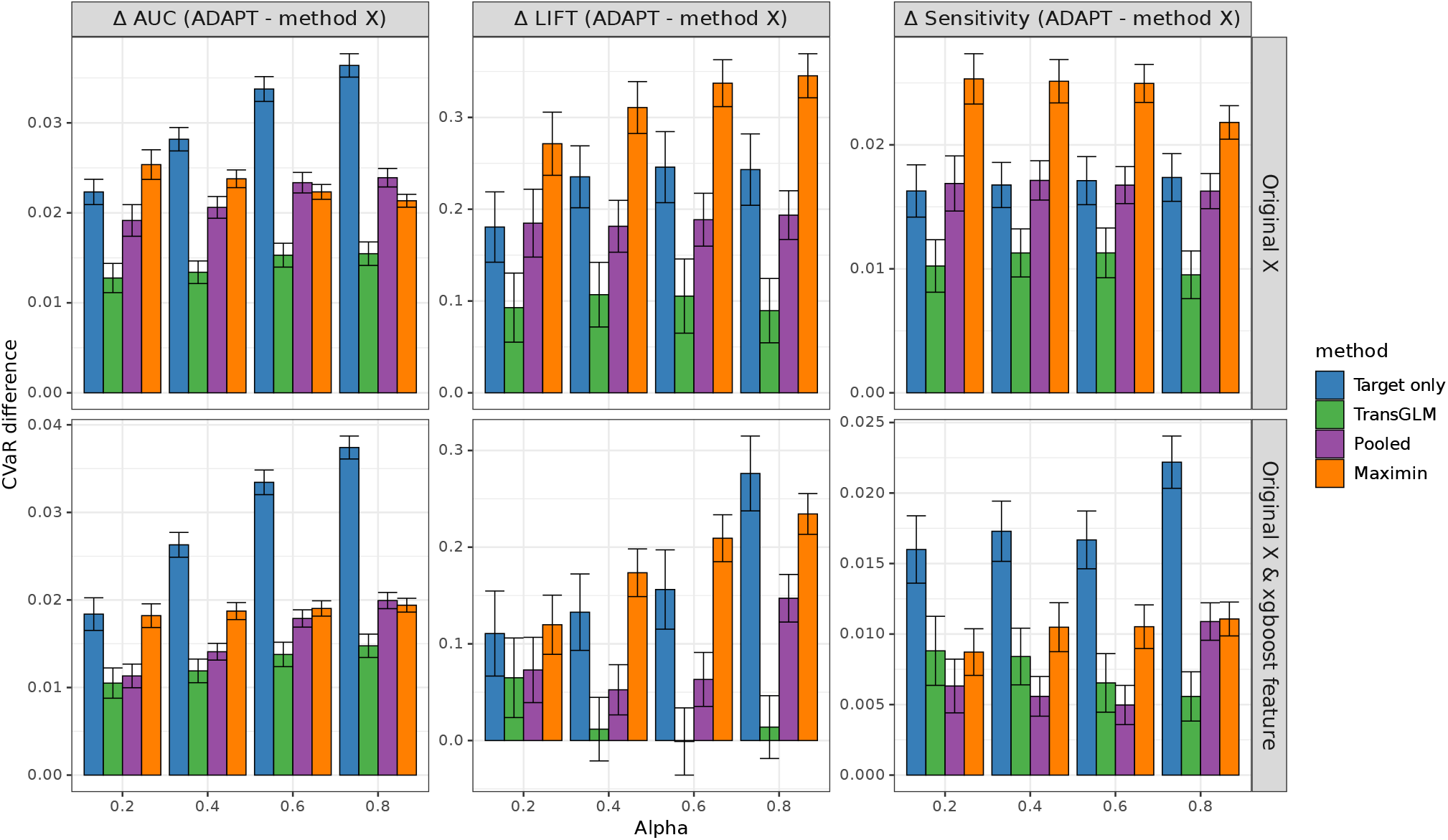
Sensitivity analysis: effect of nonlinear feature augmentation within Duke. For each benchmark method, bars show the difference in lower-tail CVaR between the benchmark and ADAPT 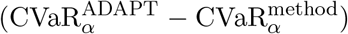, averaged across mental health outcomes, at tail fractions *α* = 0.2, 0.4, 0.6, 0.8, for AUC, lift, and sensitivity in within-Duke temporal validation. Positive values indicate worse worst-case performance than ADAPT. Methods in the top panel used the original predictors, while the lower panel additionally included the XGBoost-derived prediction scores. Error bars are 95% confidence intervals from 500 paired bootstrap replicates.

**Figure S5:**
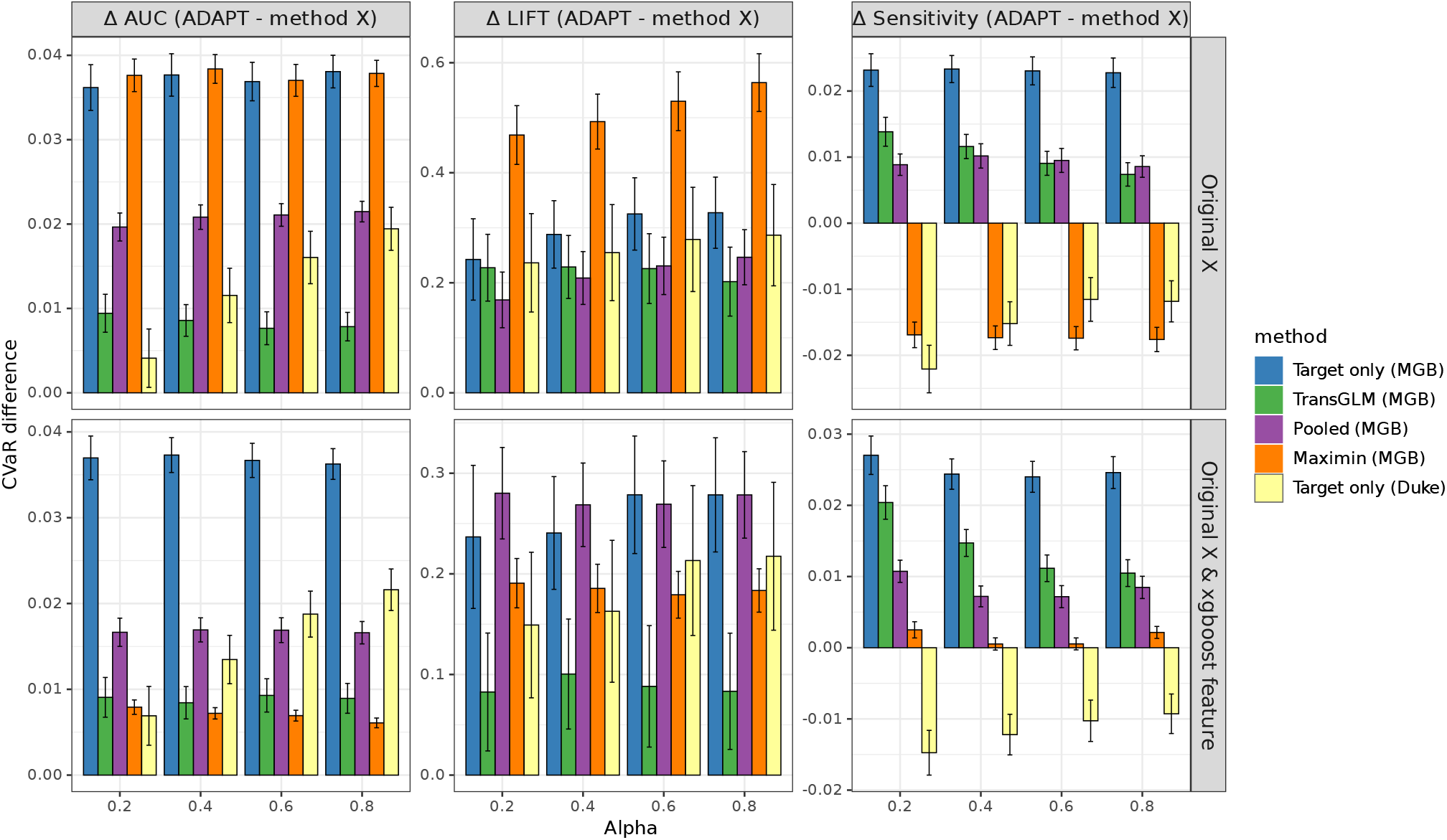
Sensitivity analysis: effect of nonlinear feature augmentation on robustness relative to ADAPT when models developed at MGB are transported to Duke. For each benchmark method, bars show the difference in lower-tail CVaR between the benchmark and ADAPT 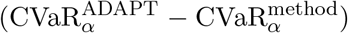, averaged across mental health outcomes, at tail fractions *α* = 0.2, 0.4, 0.6, 0.8, for AUC, lift, and sensitivity. ADAPT and the MGB benchmarks were developed at MGB and evaluated on Duke data; the Duke target-only model was developed and evaluated at Duke and serves as a local reference. Positive values indicate worse worst-case performance than transported ADAPT. Methods in the top panel used the original predictors, while the lower panel additionally included the XGBoost-derived prediction scores. Error bars are 95% confidence intervals from 500 paired bootstrap replicates.

**Figure S6:**
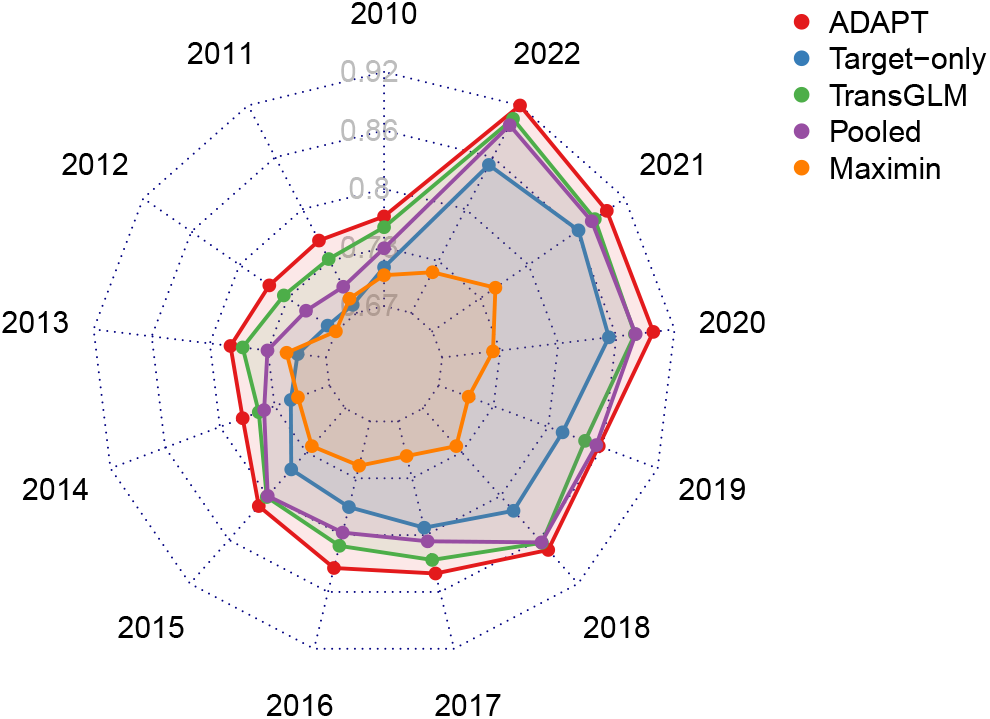
Sensitivity analysis: contemporaneous discrimination with XGBoost-augmented predictors. Suicide-attempt case study at MGB. Models were developed for a target calendar year using the original EHR predictors augmented with XGBoost prediction scores, and evaluated in the same calendar year. Points show mean AUC across repeated splits.

**Figure S7:**
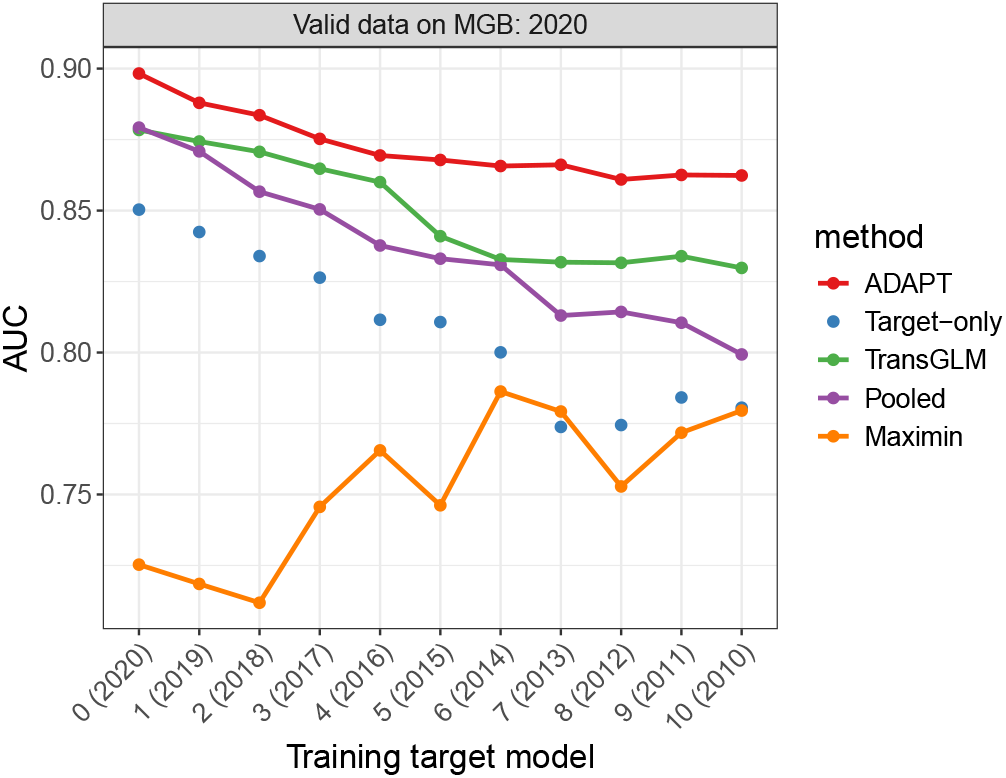
Sensitivity analysis: temporal validation in a common 2020 evaluation population with XGBoost-augmented predictors. Suicide-attempt case study at MGB. Models were developed for progressively earlier target years using the original EHR predictors augmented with XG-Boost prediction scores, and evaluated in MGB 2020 data.

**Figure S8:**
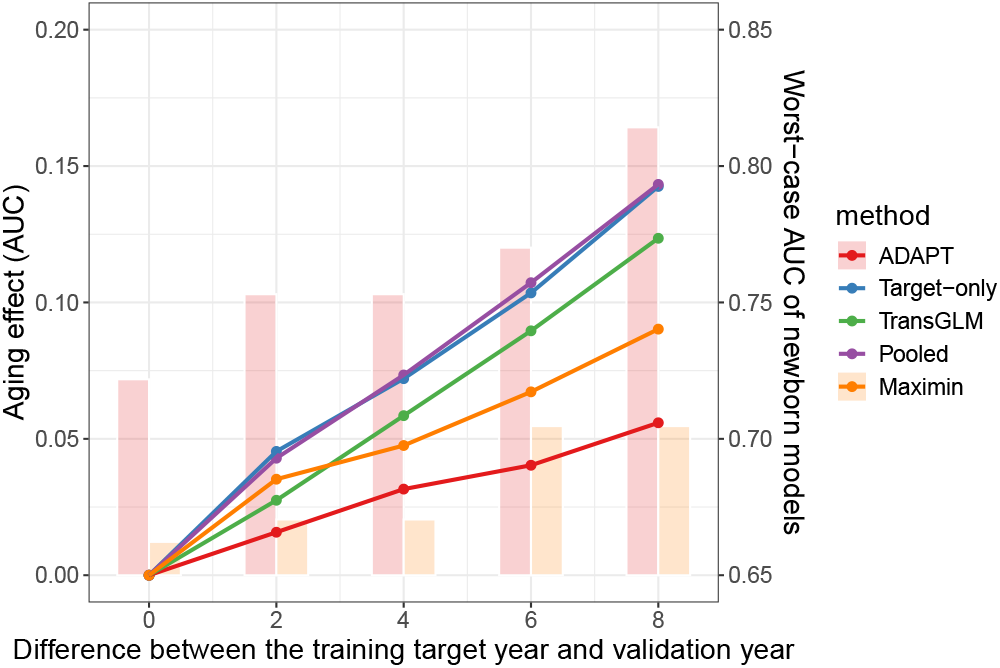
Sensitivity analysis: relative temporal degradation with XGBoost-augmented predictors. Suicide-attempt case study at MGB. Relative temporal degradation as the interval between model development and evaluation increased, for temporal validation at MGB with the original HER predictors augmented with year-specific XGBoost prediction scores. Bars show the worst (“newborn”) AUC within each interval window for ADAPT and maximin.

## Notes

### Competing Interest Statement

The authors have declared no competing interest.

### Author Declarations

Ethics committees of Duke University and Massachusetts General Hospital gave ethical approval for this work.

